# Dual methylation and hydroxymethylation study of alcohol use disorder

**DOI:** 10.1101/2020.09.16.20194639

**Authors:** Shaunna L. Clark, Robin F. Chan, Min Zhao, Lin Y. Xie, William E. Copeland, Brenda W.J.H. Penninx, Karolina A. Aberg, Edwin J.C.G. van den Oord

**Author notes:** Corresponding Author: Shaunna L. Clark; Address: 8441 Riverside Parkway, Bryan, TX, 77807.

## Abstract

Using an integrative, multi-tissue design we sought to characterize methylation and hydroxymethylation changes in blood and brain associated with alcohol use disorder (AUD). First, we used epigenomic deconvolution to perform cell-type specific methylome-wide association studies within subpopulations of granulocytes/T-cells/B-cells/monocytes in 1,132 blood samples. Blood findings were then examined for overlap with AUD-related associations in methylation and hydroxymethylation in 50 human post-mortem brain samples. Follow-up analyses investigated if overlapping findings mediated AUD-associated transcription changes in the same brain samples. Lastly, we replicated our blood findings in an independent sample of 412 individuals and aimed to replicate published alcohol methylation findings using our results.

Cell-type specific analyses in blood identified methylome-wide significant associations in monocytes and T-cells. The monocyte findings were significantly enriched for AUD-related methylation and hydroxymethylation in brain. Hydroxymethylation in specific sites mediated AUD-associated transcription in the same brain samples. As part of the most comprehensive methylation study of AUD to date, this work involved the first cell-type specific methylation study of AUD conducted in blood, identifying and replicating a finding in *DLGAP1* that may be involved in AUD-related brain impairment. In this first study to consider the role of hydroxymethylation in AUD, we found evidence for a novel mechanism for cognitive deficits associated with AUD. Our results suggest promising new avenues for AUD research.

## INTRODUCTION

Alcohol use disorder (AUD) is a devastating illness characterized by excessive, uncontrolled consumption of alcohol despite its negative consequences on the drinker’s health and well-being. DNA methylation (DNAm) studies of AUD offer unique opportunities to better understand AUD. For example, because drug-induced epigenetic changes can persist over time and have lasting effects (1), DNAm could be a potential mechanism behind phenomena such as sensitization and tolerance. Advances in DNAm array-based assays have made it possible to investigate the relationship between DNAm and AUD for hundreds of thousands of sites. While a few studies of AUD exist(2-8), they have not yet identified replicating sites across studies. This is likely due to small sample sizes, use of earlier arrays that captured a small number of sites, and inconsistent quality control, data analysis, and interpretation of significant findings(4). The one exception is a study conducted in whole blood that identified several sites associated with AUD across multiple cohorts (4). In contrast, studies of other alcohol outcomes including consumption and phosphatidylethanol, a metabolite of ethanol, have identified 7 sites that replicate across studies (9-11).

DNAm studies are typically conducted using bulk tissue (e.g., whole blood or brain) that is composed of multiple cell-types, each with a potentially different DNAm profile. In studies using bulk tissue, including some previous alcohol studies (4, 5, 9-11), estimates of cell-type proportions are included as covariates to protect against false positive associations (12). By focusing only on bulk tissue, however, potentially important associations may be missed. For example, when effects involve a single cell-type or have opposite directions in different cell-types, they may be diluted or potentially nullify each other in bulk tissue. Further, as the most common cell-types will drive the results in bulk tissue, effects from low abundance cells may be missed altogether. Additionally, knowing which specific cells harbor effects is key for the biological interpretation and crucial for designing follow-up experiments.

One way to obtain cell-type specific methylation results are to sort samples into the respective cell populations and then generate methylation data in each of the sorted cell populations. It is, however, infeasible to sort and assay methylation in each cell-type for every sample in large-scale studies. An alternative is to use a deconvolution approach which uses reference methylomes from sorted cells to test cell-type specific case-control differences. This approach has been validated for methylation data (13) and successfully applied to, for example, a cell-type specific methylation study of depression (14).

While alcohol impacts many parts of the body, DNAm studies aimed at understanding the pathogenic processes of AUD are ideally performed in brain. The brain methylome, in contrast to blood and other tissues, is more complex as hydroxymethylation (hmCG) is also common. hmCG involves oxidation of the methyl-group base of a DNA cytosine by the ten-eleven translocases proteins (TET1, TET2, TET3), predominantly in the CG context(15).

Compared to mCG, hmCG shows distinctive developmental patterns(16) and broadly impacts brain function and neural plasticity(16). Hydroxymethylation has not previously been investigated in psychiatric conditions with the exception of one study showing hydroxymethylation differences in the prefrontal cortex of depressed patients (17). While no previous study has examined AUD-related hydroxymethylation, studies have demonstrated differential expression of *TET1* in AUD(18). Studies of other drugs of abuse have found hydroxymethylation to be associated with cocaine and methamphetamine administration (19, 20). Taken together this evidence suggests that hydroxymethylation may be involved in AUD.

One limitation of working with brain tissue is it cannot be obtained from living patients. It can be collected postmortem, but obtaining the sample sizes needed for adequate statistical power is challenging. There is some research indicating methylation may be concordant across tissues at specific sites. For example, studies of the glucocorticoid receptor have shown mCG changes in both blood (21) and postmortem brain samples (22). While less studied than mCG, recent research indicates an overlap between mcG in blood and hmCG in brain in a site-specific manner (23). Multiple factors may contribute to this overlap. Peripheral tissues such as blood may reveal marks predating or resulting from the epigenetic reprogramming events affecting the germ line and embryogenesis (24), sequence variants can affect (hydroxy)methylation levels and will be identical across tissues, and systemic AUD disease processes such as inflammation will affect both blood and brain. Further, as substantial correlations have been reported between different methylation types over short distances(25), it is possible that mCG captures part of the hmCG signal. This observed overlap suggests that one approach to improve statistical power of studies of postmortem brain, is to combine results with studies in (antemortem) blood samples that are easier to collect in large sample sizes.

Our integrative design involving multiple methylation types and tissues improves statistical power and helps avoid potential study specific confounders. In the discovery, we sought to identify AUD-linked cell-type specific (granulocytes/T-cells/B-cells/monocytes) methylation (mCG; DNA methylation occurring at CpG sites) changes in blood samples in a large discovery sample of 1,132 subjects. Second, we examined if corresponding AUD-related changes can be observed in the prefrontal cortex (PFC) of AUD cases and controls from 50 independent subjects. We considered both mCG and hmCG in our brain investigations to provide a more comprehensive view of the AUD brain methylome than previous studies. Next, for AUD-associated changes that overlapped across blood and brain, we followed-up findings by studying whether the observed mCG or hmCG changes mediated case-control differences in transcript abundance levels in the same brain samples. Finally, we replicated our blood findings from the discovery in an independent sample and attempted to replicate published alcohol mCG findings. To our knowledge, this is the most comprehensive alcohol methylation study as it considers cell-type specific mCG in blood, is the largest study of AUD methylation both in terms of sample size and the number of methylation sites investigated, and considers mCG, hmCG and transcript abundance in brain from the same samples in AUD.

## MATERIALS AND METHODS

### Samples

#### Discovery Blood Samples

The mCG data was previously generated using methyl-CG binding domain sequencing (MBD-seq) (26, 27) for DNA extracted from blood samples from 1,132 individuals from the Netherlands Study of Depression and Anxiety (NESDA) (28). Lifetime AUD was diagnosed using the DSM-IV based Composite International Diagnostic Interview (CIDI version 2.1) that was administered by specially trained research staff. AUD cases (n=323) included individuals that met criteria for either abuse or dependence on the DSM-IV to match the AUD definition in the brain samples. Controls (n=809) had initiated alcohol use but did not qualify for an AUD diagnosis. The study was approved by ethical committees of all participating locations and participants provided written informed consent. Further details about the study sample, and the demographic and clinical characteristics of participants used for the present study are in Table S1.

#### Postmortem Brain Samples

The brain samples consist of 50 brain autopsies provided by the New South Wales Brain Tissue Resource Centre, Australia. Specifically, the post-mortem tissue samples are from the Brodmann Area 10 (BA10) of the PFC from 25 AUD cases and 25 unaffected controls matched to the cases on age and sex. The AUD cases consumed ≥80g alcohol per day during the majority of their adult lives and met criteria for a diagnosis of either abuse (n=10) or dependence (n=15) on the DSM-IV and AUD on DSM-5. AUD cases did not have liver cirrhosis, Wernicke–Korsakoff’s syndrome, a history of abuse of other substances beyond alcohol or major psychiatric diagnosis. Control subjects had either abstained from alcohol completely or were social drinkers who consumed ≤20g of alcohol per day on average that had no previous history of abuse of any substance or major psychiatric diagnosis. All subjects were of European descent. Further details about the subjects in the present study are in Table S2. From each tissue sample we performed simultaneous extraction of DNA and RNA using the AllPrep Universal kit (Qiagen). One aliquot of DNA was used to assay mCG and another to assay hmCG. The mCG, hmCG and transcript expression assays were all performed on the same brain tissue sample.

### Methylation and Hydroxymethylation

#### Assaying mCG

The same MBD-seq approach, which was used to generate mCG data for the blood samples (28) was also used to generate mCG data for the brain samples. Specifically, we used an optimized protocol for the MBD-seq approach (26, 27) that achieves near-complete coverage of the 28 million possible mCG sites at a cost comparable to commonly used methylation arrays (29). Briefly, genomic DNA was sheared into, on average, 150 bp fragments using the Covaris™ E220 focused ultrasonicator system. We performed enrichment with MethylMiner™ (Invitrogen) to capture the mCG fraction of the genome. MethylMiner is specific for mCG (29), and, in contrast to bisulfite-based approaches, MethylMiner does not detect hydroxymethylation. Next, dual-indexed sequencing library for each methylation capture was prepared using the Accel-NGS® 2S Plus DNA Library Kit (Swift Biosciences). Ten libraries were pooled in equal molarities and sequenced with 75 cycles (i.e., 1 × 75 base pair reads) on a NextSeq500 instrument (Illumina).

#### Assaying hmCG

As hydroxymethylation is very rare in blood, hmCG was only assayed in the brain samples. To assay hmCG, selective chemical labeling and enrichment of hmC (hMe-Seal) was performed using components of the Hydroxymethyl Collector kit (Active Motif). We substituted the enzyme in the kit with T4 β-glucosyltransferase (New England Biosciences #M0357) to improve labeling performance. In our experience, using high-quality and high-activity enzyme is critical for successful hmC enrichment. T4 β-glucosyltransferase was used to selectively label hmC residues with 150 μM final UDP-azide-glucose. Each azo-glucosylated hmC was biotinylated via dibenzocyclooctyl click chemistry by addition of Biotin Conjugate Solution. The labeled DNA was then column purified, enriched with paramagnetic streptavidin beads, washed and eluted following the vendor protocol, with the substitution of end-over-end rotation with agitation on an orbital shaker at 700 rpm in a 96-well 1.2 ml square well plate. Next, dual-indexed sequencing library for each hydroxymethylation capture was prepared using the Accel-NGS® 2S Plus DNA Library Kit (Swift Biosciences). Ten libraries were pooled in equal molarities and sequenced with 75 cycles (i.e., 1 × 75 base pair reads) on a NextSeq500 instrument (Illumina).

#### mCG and hmCG Data Processing

The sequence reads were aligned to the human reference genome (hg19/GRCh37) using Bowtie2 (30). Data quality control and analyses were performed in RaMWAS (31). mCG and hmCG scores were calculated by estimating the number of fragments covering each site using a non-parametric estimate of the fragment size distribution (31). These scores provide a quantitative measure of mCG/hmCG for each individual at that specific site (29). See supplemental material for further details.

#### Methylome-wide association testing (MWAS)

MWAS testing in blood was performed with RaMWAS (31) using multiple regression analyses that included several classes of covariates. These classes were measured technical variables such as batch and peak (31), demographic variables for age and sex, and estimated cell-type proportions (12). We also included smoking (yes/no; current use of cigarettes) and depressive disorder (yes/no; DSM-IV-based diagnosis) status as covariates. Principal component analysis was performed on the covariate-adjusted methylation data to capture any remaining unmeasured sources of variation. We used a Scree test to select three principal components to include in the final blood MWAS.

To account for multiple testing, we used a false discover rate approach (FDR). Unlike methods that control the family-wise error rate (e.g., the Bonferroni correction), the FDR is robust against having correlated tests because it controls a ratio. Furthermore, FDR control does not depend on the number of tests (it depends on the proportion of tests without effects). To declare methylome-wide significance, we applied a FDR threshold of 0.1. Operationally, the FDR was controlled using *q*-values that are FDRs calculated using the *p*-values of the individual tests as thresholds for declaring significance. We chose a q-value of 0.1 as the threshold because more stringent thresholds result in exponential decreases in power (32).

The brain mCG and hmCG association testing was conducted separately for mCG and hmCG, and included measured technical variables, sex, age at death, post-mortem interval, estimated cell-type proportions, and one principal component. Smoking and depression status were not included as covariates in the brain association testing due to this information being unavailable.

#### Cell-type Specific MWAS

We used an epigenomic deconvolution approach to perform cell-type specific MWAS for the major nucleated cell-types in blood: T-cells (CD3+), monocytes (CD14+), granulocytes (CD15+) and B-cells (CD19+). This approach applies statistical methods in combination with MBD-seq mCG profiles from a reference set of purified cells to deconvolute the cell-type specific effects from data generated in bulk tissue. A description of this approach and the generation of the reference panels has previously been described elsewhere (14). In short, cell-type proportion estimates for each sample were obtained using Houseman’s method (12). These estimates were used to test the null hypothesis that methylation of a given site is not correlated with AUD status for each cell-type (33). Cell-type proportions were included as main effects in the cell-type specific MWAS as the epigenomic deconvolution is essentially an interaction model(33) (see Supplemental Material). To declare methylome-wide significance we applied a FDR threshold of 0.1. Deconvolution was not applied to the brain samples as the deconvolution approach relies on an interaction effect to detect the cell-type specific case-control differences. With only 50 samples, the deconvolution analyses would be underpowered.

#### Pathway Analysis

To gain insight into the biological pathways affected by AUD, we used ConsensusPathDB (CPDB) to test for overrepresentation of top brain findings (*P<*1×10^−5^) located within genes in the biological pathways in the Reactome database. A minimum of three genes from the top findings had to be present in the pathway for it to be considered enriched.

#### Cross-tissue Overlap and Colocalization Testing

To explore if there is overlapping AUD-associated methylation at the same sites across blood and brain, we tested whether the blood mCG findings were enriched for brain mCG and hmCG top findings. Specifically, testing was conducted using circular permutations that generate the empirical test-statistic distribution under the null hypothesis while preserving the correlational structure of the data. We defined the “top” findings as the top 0.1% and 0.5% of all sites tested from our MWASs based on their p-values with the lowest 0.1% and 0.5% of p-values constituting the “top” and anything below those thresholds constituting the bottom. This corresponds to 21,868 and 109,342 findings in the top 0.1% and 0.5% for blood, 20,823 and 104,117 for brain mCG, and 26,153 and 130,769 for brain hmCG, respectively. We corrected for testing multiple thresholds by using the same threshold in the permutations used to generate the null distribution. We applied the same approach to test whether the overlapping sites across tissues colocalized with UCSC Genome Browser genomic feature tracks and Roadmap Epigenomics Project chromHMM 15-state chromatin model tracks (34). Further details can be found in the Supplemental Material.

### Transcript Expression

#### Assaying Transcript Abundance

We used high-quality RNA extracted from the brain tissue samples, with an average RNA integrity number (RIN) of 9.3 (sd=0.58), to assess transcript abundance with RNA-seq. For each sample 800 ng of total RNA was used with 0.8 reactions of the TruSeq Stranded Total RNA Gold sample preparation kit (Illumina) to perform bead-based depletion of cytoplasmic and mitochondrial rRNA, followed by cDNA synthesis. Using Truseq RNA UD indexes (Illumina), amplification was performed to generate a uniquely labeled paired-end sequencing library for each sample. Following library preparation, appropriate library size was confirmed using the Agilent 2100 Bioanalyzer with the high sensitivity DNA kit (Agilent) and fluorometric quantification of the concentration of double stranded DNA was assessed via Qubit (ThermoFisher Scientific). Eight libraries were pooled in equal molarities and sequenced with 150 cycles (i.e., 2 × 75 base pair reads) on a NextSeq500 instrument (Illumina).

Transcriptome sequence reads were aligned using HiSat2 (35). Transcript assembly and estimation of abundance levels per transcript were performed using StringTie (36). Abundance levels were normalized to transcript per million (TPM). Further details are provided in the supplemental material.

#### Testing Methylation Mediation

We used the transcript abundance levels to examine if mCG/hmCG sites that showed case-control differences across tissues were related to AUD-associated transcription changes. Specifically, for mCG sites in cell-types that showed differential methylation with AUD in blood, we tested whether the mCG\hmCG levels in brain of sites that overlapped across tissues were associated with transcript abundance (i.e., cis-meQTL) in the same brain samples. To declare a site a potential cis-meQTL, we used a p-value threshold of 0.01. Mediation analyses were then conducted with the significant cis-meQTLs to determine if the AUD-associated variation in mCG/hmCG accounted for possible case-control differences related to AUD-associated transcription changes. Mediation testing was conducted with the mediate function of the mediation R package using a quasi-Bayesian approach with 100,000 Monte Carlo draws for the approximation of the p-values for the mediation effect (37). A Bonferroni-correction was used to declare significant mediation.

### Replication

#### Replication of blood mCG findings in an independent sample

The mCG data for the replication comes from an adult subsample of 412 individuals from the Great Smoky Mount Study (GSMS; (38)). The mCG data was generated using the same MBD-seq approach as the discovery for DNA extracted from blood spots. AUD was diagnosed using the Young Adult Psychiatric Assessment (YAPA). There were 73 AUD cases and 339 controls that had initiated alcohol use but did not qualify for an AUD. Further details about the replication sample participants, and mCG data generation, quality control and analysis are described in the Supplement Material, but generally follow the same procedure used for the discovery.

To replicate our blood findings, we first determined if there were overlapping sites between the discovery and replication by testing if the replication findings were enriched for the discovery findings. This was done using the circular permutation testing described above using the same thresholds of 0.1% and 0.5% to define the “top” findings. This corresponds to 22,670 and 113,353 findings being in the top 0.1% and 0.5% for the replication. For cell-types that showed significant enrichment, we then performed a look-up replication of the significant findings from the discovery. A site was considered replicated if it had the same direction of effect as the discovery and the p-value was less than the Bonferroni adjusted p-value threshold for the number of tests performed per cell-type. A site was considered nominally replicated if it had the same direction of effect and p-value<0.05.

#### Replication of existing mCG findings

To replicate existing alcohol mCG findings, we compiled a list of published array-based alcohol mCG studies (Table S3). Only a few of these studies reported sufficient information about their significant findings to allow for replication attempts like the location of their significant findings (i.e., chromosome and base-pair position) or the probe identification number (i.e., cg-number). For studies that reported sufficient information, we performed a look-up replication in our mCG data in whole blood and brain on all reported significant individual CpGs and CpGs located in significant differentially methylated regions. Replication was established using the same criteria as in the independent sample.

## RESULTS

In total, 21,868,402 mCGs were investigated in blood, 20,823,597 mCGs and 26,153,809 hmCGs were investigated in the brain tissue. The genomic distribution of methylated sites in blood and brain, and hydroxymethylated sites in brain is shown in Figure S1.

### Discovery: Blood MWAS

The Quantile-Quantile (QQ) plots for whole blood and individual cell-types suggested no test statistic inflation with lambda (ratio of the median of the observed distribution of the test statistic to the expected median) ranging from 0.97-1.06 (Figure 1). Methylome-wide significant findings were detected for monocytes and T-cells but not for whole blood, granulocytes or B-cells (Tables S4-S8). In T-cells, 3 CpGs passed methylome-wide significance with the top two sites (p=3.72×10^−10^ and 1.24×10^−09^) found in the long intergenic noncoding RNA *RP11-342D11*.*3* (Table S8). The MWAS for monocytes yielded 1,397 methylome-wide significant CpGs (FDR<0.1; Table S6), with the top genic findings located in *BAZ2B* and *PLA2G4A*.

**Figure 1.**
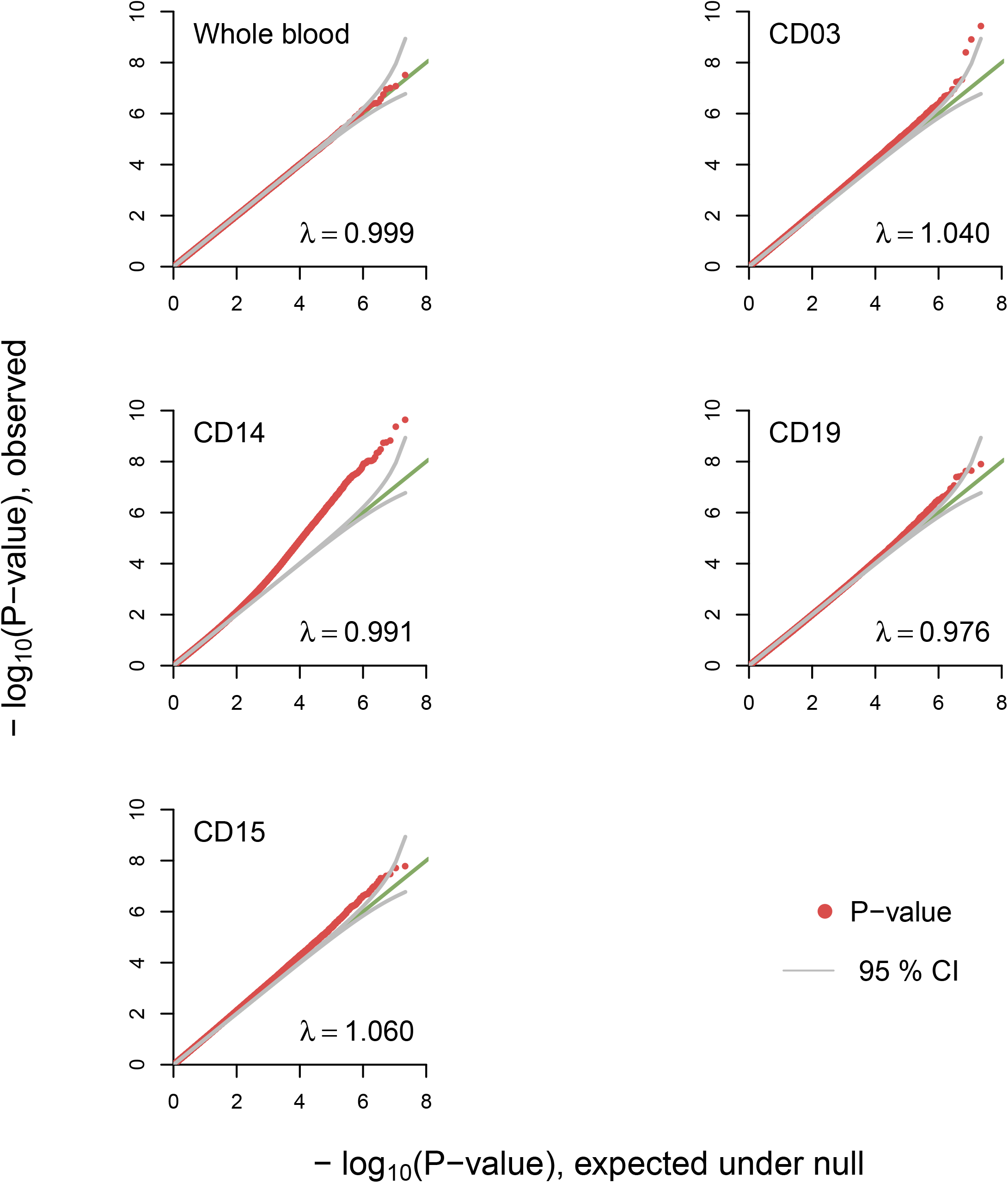
Quantile-Quantile plots for whole blood and cell-type specific MWAS of AUD The observed p-values (red dots), on a -log10 scale, are plotted against their expected values (grey main diagonal line) under the null hypothesis assuming none of the CpGs have an effect. Grey lines indicate the 95% confidence bands (CI). A deviation of the observed p-values from the main diagonal indicates that there are CpGs associated with AUD. Coefficient lambda (λ) will be close to one if the vast majority of CpGs behave as expected under the null hypothesis. Analyses were performed in whole blood as well as blood cell subpopulations using a reference panel that was sorted using cell surface antigen molecules CD3, CD14, CD15, and CD19 that are expressed on the surface of T-cells, monocytes, granulocytes, and B-cells, respectively.

### Brain Overlap

The exploratory methylome-wide results for both mCG and hmCG showed that no individual site reached methylome-wide significance in brain (Table S9 and S10). As a supplemental analysis, we combined proximal mCG and hmCG sites, respectively, to form “blocks” and then tested these “blocks” for association (see Supplemental Material). This analysis generated similar results as no block reached significance in either mCG or hmCG (Figure S2 & S3). The top mCG finding was located in *GOSR2* (p=3.98×10^−08^), a member of the golgi SNAP receptor complex gene family linked to synaptic integrity(39). The top hmCG finding was located in the spastin gene (*SPAST*; p=1.54×10^−07^) which has been linked to vesicle trafficking in neurons(40). Pathway analyses revealed the potential for mCG to alter pathways related to the immune system and G-protein coupled receptors by hmCG (Table S11).

To assess whether there was overlapping AUD-related methylation across blood and brain, we tested whether the top whole blood and cell-type specific MWAS results were enriched for the top mCG and hmCG results in brain. We observed significant overlap (Table 1) in top sites between the monocyte MWAS with both mCG (2,315 sites) and hmCG (2,366 sites) in brain and the B-cell MWAS with both mCG (144 sites) and hmCG (38 sites).

**Table 1.** Enrichment of Top Blood MWAS findings for brain MWAS findings

| Blood MWAS | Brain Methylation |  |  |  | Brain Hydroxymethylation |  |  |  |
| --- | --- | --- | --- | --- | --- | --- | --- | --- |
|  | Gene Overlap | Top Threshold | Odds Ratio | P-value | Gene Overlap | Top Threshold | Odds Ratio | P-value |
| Whole Blood | 525 | 0.5;0.5 | 1.019 | 0.667 | 97 | 0.5;0.1 | 0.883 | 0.987 |
| T-cells (CD03+) | 98 | 0.5;0.1 | 0.997 | 0.814 | 99 | 0.5;0.1 | 0.905 | 0.977 |
| Monocytes (CD14+) | 149 | 0.1;0.5 | 1.378 | 0.009 | 615 | 0.5;0.5 | 1.128 | 0.035 |
| Granulocytes (CD15+) | 96 | 0.1;0.5 | 0.982 | 0.858 | 108 | 0.1;0.5 | 0.988 | 0.846 |
| B-cells (CD19+) | 144 | 0.1;0.5 | 1.368 | 0.007 | 38 | 0.1;0.1 | 1.741 | 0.011 |

MWAS, methylome-wide association study; Top Threshold, first number reflects the percentage of top results from blood and the second number reflects the percentage of top results from the brain.

### Characterization

#### Colocalization of Overlap

Focusing on the monocytes, which showed methylome-wide significant findings in blood and had significant overlap with the brain results, we characterized the overlap by testing if the overlapping sites were enriched for specific genomic features and chromatin states (Tables S12 & S13). The overlap between monocytes and both mCG and hmCG was enriched at CpG islands, exons, genes, gene promotors and DNase hypersensitivity regions. Both the mCG and hmCG overlap with monocytes were enriched for the enhancer chromatin state.

#### Mediation testing of cis-MeQTLs with brain expression data

mCG and hmCG findings that overlapped between blood and brain tissue were tested for association with transcript levels (i.e., cis-meQTL testing) in brain. Of the overlapping sites, two were associated with transcript level expression for monocyte/mCG and six for monocyte/hmCG (Table 2).

**Table 2.** Significant cis-MeQTLs and mediation results

| Methylation Type | Gene | Chr | BP | Transcript Name | MeQTL Effect | MeQTL p-value | Mediation Effect | Mediation P-value |
| --- | --- | --- | --- | --- | --- | --- | --- | --- |
| hmCG | *ANKRD13C* | 1 | 70775032 | ENST00000498735 | -0.129 | 0.004 | -0.067 | 0.624 |
| hmCG | *ANKRD13C* | 1 | 70775035 | ENST00000498735 | -0.127 | 0.004 | -0.071 | 0.596 |
| mCG | *ERC2* | 3 | 56104924 | ENST00000466358 | 0.117 | 0.007 | 0.065 | 0.672 |
| hmCG | *ERICH1-AS1* | 8 | 843888 | ENST00000524139 | 0.151 | 0.007 | 0.065 | 0.534 |
| hmCG | *PRMT7* | 16 | 68350898 | ENST00000441236 | 0.161 | 0.002 | 0.042 | 0.730 |
| hmCG | *BAIAP2* | 17 | 79062899 | ENST00000572073 | -0.165 | 0.002 | -0.407 | 0.0014 |
| hmCG | *NEDD4L* | 18 | 55923590 | ENST00000590248 | -0.142 | 0.002 | -0.087 | 0.558 |
| mCG | *NFATC1* | 18 | 77193667 | ENST00000545796 | 0.119 | 0.009 | 0.268 | 0.266 |

For the significant cis-meQTLs, we performed mediation analyses to test whether mCG/hmCG mediated the relationship between AUD and transcript expression. There was a significant mediation effect after Bonferroni correction (α=0.05/8=0.00625; Table 2) between hmCG at a site in *BAIAP2*, ENST00000572073, a transcript of *BAIAP2*. The regression coefficients between AUD and transcript expression and hmCG level were both significant (Figure 2). We observed a significant mediation effect (p=0.00142) where the mediation effect was 2.09×-0.19 = -0.40. As the regression coefficient between AUD and expression is significant in the mediation model, it suggests that hmCG partially mediates the relationship between AUD and transcript expression at this location.

**Figure 2.**
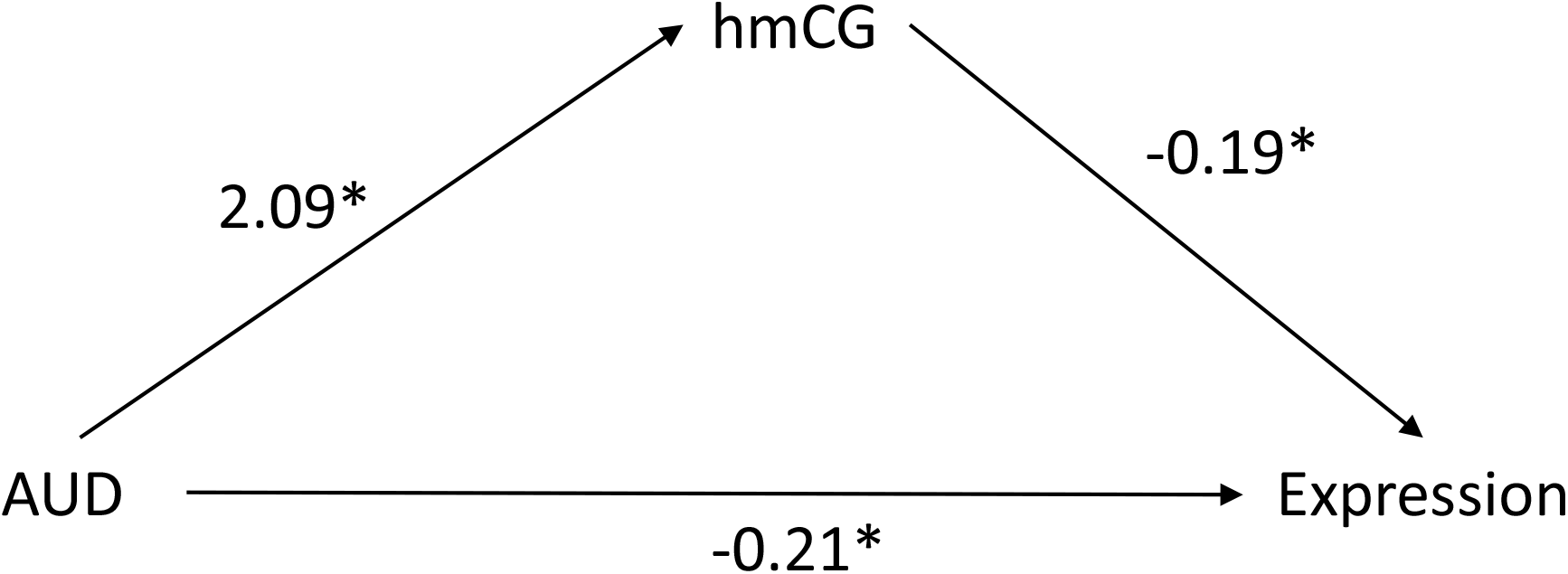
Unstandardized regression coefficients for the relationship between AUD and expression of transcript ENST00000572073 as mediated by hydroxymethylation (hmCG). *p < 0.05; hmCG is the hydroxymethylation level at location 79,062,899 on chromosome 17

### Replication

#### Replication of blood mCG findings in an independent sample

We observed significant overlap (Table S16) in top sites between the monocyte MWAS in the discovery and replication (80 sites, p<1×10^−06^) and B-cells (66 sites, p<1×10^−06^). The look-up replication yielded one replicating site in *DLGAP1* in monocytes (p=5.30×10^−05^) after multiple testing correction and an additional 34 sites that nominally replicated.

#### Replication of existing mCG findings

To replicate existing mCG findings from bulk tissue, we performed a look-up replication in our whole blood and brain mCG results (Tables S14 and S15). We were able to replicate one finding at cg18159646 (chr 11; bp=6,262,1179) in *SNORD22/SNHG1* (4) after Bonferonni correction and an additional nominally significant finding (Table S14 bolded). Both of these sites had the same direction of effect as our study. In brain, we were unable to replicate any sites after multiple testing correction, however were were able to replicate 27 findings at a nominal threshold with the same direction of effect.

## DISCUSSION

In one of the most comprehensive methylation studies of AUD to date, we identified several novel findings that may provide potential mechanisms behind the cognitive and other brain-related impairments seen in AUD patients. Of interest is a replicating finding from monocytes in *DLGAP1* (DLG associated protein 1) which influences postsynaptic density through its involvement in the DLGAP1-DLG4-NMDA pathway. Disruption of this pathway at the *DLGAP1* level leads to physical impairments of postsynaptic density organization(41), which may play a role in the cognitive and social deficits seen in some psychiatric disorders(42, 43).

While our replicating finding was in blood, mCG sites in *DLGAP1* in our brain MWAS were significant (p<1×10^−5^) and among the cross-tissue overlap. mCG in *DLGAP2*, a member of same gene family, was previously associated with AUD and showed a similar pattern of overlapping methylation between blood and brain(5). This suggests that our blood finding may mirror a potential process in brain whereby AUD-associated mCG in *DLGAP1* disrupts the DLGAP1-DLG4-NMDA pathway leading to changes in postsynaptic density and downstream brain impairments.

We also aimed to replicate findings from existing mCG studies. This effort was a modest success as we were able to replicate one finding in blood after multiple testing correction, and one additional finding in blood and 27 in brain at a nominal threshold. There are several reasons why we see replication of a few sites. First, for the brain studies, including our own, the sample sizes were 50 or less which may only have modest power to detect effects. Second, some of the previous studies did not control for cell-type heterogeneity, which can induce false positive results (12). However, most of these studies were published before it became standard practice to control for cell-type heterogeneity. Third, analyses conducted using methylation arrays can be sensitive to quality control procedures used, and unless stringent quality control is applied array-based methylation studies can show an excess false positive findings due to test statistic inflation (44). The level of inflation can be assessed through lambda (the median of the observed test statistics divided by the expected median of the test statistics under the assumed null distribution) with values close to 1 indicating a lack of inflation. Unfortunately, lambda was not reported for the methylation array studies we attempted to replicate and it is therefore hard to assess whether this would potentially explain the replication of only a few sites.

We also examined if changes in blood mCG showed corresponding AUD-related mCG and hmCG changes in brain tissue. The cross-tissue analyses indicated significant overlapping AUD-related methylation signals across blood and brain at specific sites. This finding aligns with previous alcohol mCG studies in both model organisms and humans showing there are overlapping methylation sites associated with alcohol across whole blood and bulk brain tissue (2, 5, 45). It should be noted that overlapping sites in blood and brain, both in our study and the previous studies, did not always have the same direction of effect likely due to alcohol having a different impact in each tissue. As we explored the overlap between blood mCG and brain hmCG we would not expect the direction of effect to be the same. Overall, this overlap across tissues has potential applications as it suggests that alcohol-related methylation changes in blood at specific sites may be biomarkers for mCG or hmCG changes occurring in brain.

A novel aspect of our study is the consideration of hydroxymethylation within the context of AUD. Our main finding with hydroxymethylation involved a site in *BAIAP2* (BAR/IMD domain

containing adaptor protein 2) where hmCG was associated with AUD, and partially mediated the relationship between AUD and abundance levels at transcript ENST00000572073. Although, the function of the specific isoform is unknown, the gene encodes for a brain-specific angiogenesis inhibitor (BAI1)-binding protein. *BAIAP2* has been linked to several psychiatric disorders (46, 47). Mice lacking *Baiap2* (i.e., *IRSp53*^*-/-*^) show cognitive and social deficits and hyperactivity, clinical features of the psychiatric disorders linked to *BAIAP2* (48). *Baiap2* is differentially expressed in the brains of alcohol exposed rats in both a binge-drinking model and a fetal alcohol exposure model (49). While our finding requires further validation, it suggest that hydroxymethylation in *BAIAP2* in AUD cases may lead to changes in expression. This change in expression of *BAIAP2* may lead to the cognitive deficits and other brain-impairments often seen in alcohol dependent patients.

Our findings must be interpreted in the context of potential limitations. All biosamples were collected after the development of AUD. Thus, some of the associated sites may have been present before AUD onset, and therefore may be susceptibility loci, while others may have occurred after AUD onset and instead reflect the disease state. To properly disentangle these effects, a study design that also included pre-AUD onset and potentially pre-drinking onset biosamples would be required. This design is not feasible for studies conducted in human postmortem brain tissue, and would only apply to studies conducted in human peripheral tissues or model organisms. Further, our results suggested potential mechanisms through which AUD-related mCG and hmCG may lead to the development of brain impairments. For findings that overlapped in brain, some sites did influence expression in brain tissue providing some evidence of functional effects. Possible next steps to test these suggested mechanisms include testing findings in independent samples and exploring the mechanism through functional experiments. For example, clustered regularly interspaced short palindromic repeats‐associated protein 9 (CRISPR‐Cas9) based technologies would enable targeted alterations to test the downstream functional effects of associated mCG/hmCG experimentally (50).

Using an integrative approach involving multiple methylation types and tissues, we were able to conduct a comprehensive investigation of AUD-related methylation. This includes an investigation of cell-type specific mCG in AUD conducted in blood that identified and replicated sites in *DLGAP1* that are potentially involved in alcohol-related brain impairment. We also found significant overlap of AUD-related mCG and hmCG on site-specific basis between blood and brain. In one of the first studies to consider hydroxymethylation in AUD, we found that hydroxymethylation in *BAIAP2* mediated AUD expression in the PFC, potentially providing a novel mechanism for AUD cognitive impairments. Our results suggest promising new avenues for AUD research as associated methylation sites have profound translational potential as methylation can be modified by drugs and targeted epigenetic editing.

## Data Availability

Data is available upon request and approval from the authors.

## Acknowledgments

This study was supported by the National Institutes of Health (R01MH099110 & 1R01MH104576 to E.J.C.G., and 1R01AA026057 to S.L.C.), the VCU CTSA (UL1TR002649 from the National Institutes of Health’s National Center for Advancing Translation Science and CCTR Endowment Fund of the Virginia Commonwealth University, and a NARSAD Young Investigator Grant from the Brain & Behavior Research Foundation. The funding sources had no role in the study design, writing of the report or decision to submit the article for publication.

The infrastructure for the NESDA study (www.nesda.nl) is funded through the Geestkracht program of the Netherlands Organisation for Health Research and Development (ZonMw, grant number 10-000-1002) and financial contributions by participating universities and mental health care organizations (VU University Medical Center, GGZ inGeest, Leiden University Medical Center, Leiden University, GGZ Rivierduinen, University Medical Center Groningen, University of Groningen, Lentis, GGZ Friesland, GGZ Drenthe, Rob Giel Onderzoekscentrum).

Tissues were received from the New South Wales Brain Tissue Resource Centre at the University of Sydney. Research reported in this publication was supported by the National Institute of Alcohol Abuse and Alcoholism of the National Institutes of Health under Award Number R28AA012725. The content is solely the responsibility of the authors and does not represent the official views of the National Institutes of Health.

## Data Availability Statement

The data that support the findings of this study are available on request from the corresponding author. Some of the data are not publicly available due to privacy or ethical restrictions. Full summary results are available upon request from the corresponding author.

## Author Acknowledgement

SC, KA, EvdO were responsible for the study design and concept. RC, MA, LX, WC, BP contributed to the acquisition of phenotypic and sequencing data, SC and EvdO performed the statistical analyses, RC, WC, BP, and KA assisted with data analysis and interpretation of study findings. SC drafted the manuscript. RC, WC, BP, KA EvdO provided critical revision of the manuscript for important intellect content. All authors critically reviewed the context and approved the final version for publication.

## REFERENCES

1. Nestler EJ. Epigenetic mechanisms of drug addiction. Neuropharmacology. 2014;76 Pt B:259–68.

2. Gatta E, Grayson DR, Auta J, Saudagar V, Dong E, Chen Y, et al. Genome-wide methylation in alcohol use disorder subjects: implications for an epigenetic regulation of the cortico-limbic glucocorticoid receptors (NR3C1). Mol Psychiatry. 2019.

3. Hagerty SL, Bidwell LC, Harlaar N, Hutchison KE. An Exploratory Association Study of Alcohol Use Disorder and DNA Methylation. Alcohol Clin Exp Res. 2016;40(8):1633–40.

4. Lohoff FW, Roy A, Jung J, Longley M, Rosoff DB, Luo A, et al. Epigenome-wide association study and multi-tissue replication of individuals with alcohol use disorder: evidence for abnormal glucocorticoid signaling pathway gene regulation. Mol Psychiatry. 2020.

5. Meng W, Sjoholm LK, Kononenko O, Tay N, Zhang D, Sarkisyan D, et al. Genotype-dependent epigenetic regulation of DLGAP2 in alcohol use and dependence. Mol Psychiatry. 2019.

6. Zhang R, Miao Q, Wang C, Zhao R, Li W, Haile CN, et al. Genome-wide DNA methylation analysis in alcohol dependence. Addict Biol. 2013;18(2):392–403.

7. Wang F, Xu H, Zhao H, Gelernter J, Zhang H. DNA co-methylation modules in postmortem prefrontal cortex tissues of European Australians with alcohol use disorders. Sci Rep. 2016;6:19430.

8. Lin H, Wang F, Rosato AJ, Farrer LA, Henderson DC, Zhang H. Prefrontal cortex eQTLs/mQTLs enriched in genetic variants associated with alcohol use disorder and other diseases. Epigenomics. 2020;12(9):789–800.

9. Liang X, Justice AC, So-Armah K, Krystal JH, Sinha R, Xu K. DNA methylation signature on phosphatidylethanol, not on self-reported alcohol consumption, predicts hazardous alcohol consumption in two distinct populations. Mol Psychiatry. 2020.

10. Xu K, Montalvo-Ortiz JL, Zhang X, Southwick SM, Krystal JH, Pietrzak RH, et al. Epigenome-Wide DNA Methylation Association Analysis Identified Novel Loci in Peripheral Cells for Alcohol Consumption Among European American Male Veterans. Alcohol Clin Exp Res. 2019;43(10):2111–21.

11. Liu C, Marioni RE, Hedman AK, Pfeiffer L, Tsai PC, Reynolds LM, et al. A DNA methylation biomarker of alcohol consumption. Mol Psychiatry. 2018;23(2):422–33.

12. Houseman EA, Accomando WP, Koestler DC, Christensen BC, Marsit CJ, Nelson HH, et al. DNA methylation arrays as surrogate measures of cell mixture distribution. BMC Bioinformatics. 2012;13:86.

13. Zheng SC, Breeze CE, Beck S, Teschendorff AE. Identification of differentially methylated cell types in epigenome-wide association studies. Nat Methods. 2018;15(12):1059–66.

14. Chan RF, Turecki G, Shabalin AA, Guintivano J, Zhao M, Xie LY, et al. Cell Type-Specific Methylome-wide Association Studies Implicate Neurotrophin and Innate Immune Signaling in Major Depressive Disorder. Biol Psychiatry. 2020;87(5):431–42.

15. Ito S, D/’Alessio AC, Taranova OV, Hong K, Sowers LC, Zhang Y. Role of Tet proteins in 5mC to 5hmC conversion, ES-cell self-renewal and inner cell mass specification. Nature. 2010;466(7310):1129–33.

16. Wang T, Pan Q, Lin L, Szulwach KE, Song CX, He C, et al. Genome-wide DNA hydroxymethylation changes are associated with neurodevelopmental genes in the developing human cerebellum. Hum Mol Genet. 2012;21(26):5500–10.

17. Gross JA, Pacis A, Chen GG, Drupals M, Lutz PE, Barreiro LB, et al. Gene-body 5-hydroxymethylation is associated with gene expression changes in the prefrontal cortex of depressed individuals. Transl Psychiatry. 2017;7(5):e1119.

18. Gatta E, Auta J, Gavin DP, Bhaumik DK, Grayson DR, Pandey SC, et al. Emerging Role of One-Carbon Metabolism and DNA Methylation Enrichment on delta-Containing GABAA Receptor Expression in the Cerebellum of Subjects with Alcohol Use Disorders (AUD). Int J Neuropsychopharmacol. 2017;20(12):1013–26.

19. Feng J, Shao N, Szulwach KE, Vialou V, Huynh J, Zhong C, et al. Role of Tet1 and 5-hydroxymethylcytosine in cocaine action. Nat Neurosci. 2015;18(4):536–44.

20. Jayanthi S, Gonzalez B, McCoy MT, Ladenheim B, Bisagno V, Cadet JL. Methamphetamine Induces TET1- and TET3-Dependent DNA Hydroxymethylation of Crh and Avp Genes in the Rat Nucleus Accumbens. Mol Neurobiol. 2018;55(6):5154–66.

21. Perroud N, Paoloni-Giacobino A, Prada P, Olie E, Salzmann A, Nicastro R, et al. Increased methylation of glucocorticoid receptor gene (NR3C1) in adults with a history of childhood maltreatment: a link with the severity and type of trauma. Transl Psychiatry. 2011;1:e59.

22. McGowan PO, Sasaki A, D’Alessio AC, Dymov S, Labonte B, Szyf M, et al. Epigenetic regulation of the glucocorticoid receptor in human brain associates with childhood abuse. Nat Neurosci. 2009;12(3):342–8.

23. Lardenoije R, Roubroeks JAY, Pishva E, Leber M, Wagner H, Iatrou A, et al. Alzheimer’s disease-associated (hydroxy)methylomic changes in the brain and blood. Clin Epigenetics. 2019;11(1):164.

24. Efstratiadis A. Parental imprinting of autosomal mammalian genes. Curr Opin Genet Dev. 1994;4(2):265–80.

25. Lee JH, Park SJ, Nakai K. Differential landscape of non-CpG methylation in embryonic stem cells and neurons caused by DNMT3s. Sci Rep. 2017;7(1):11295.

26. Aberg KA, Chan RF, Shabalin AA, Zhao M, Turecki G, Heine Staunstrup N, et al. A MBD-seq protocol for large-scale methylome-wide studies with (very) low amounts of DNA. Epigenetics. 2017:10.

27. Chan RF, Shabalin AA, Xie LY, Adkins DE, Zhao M, Turecki G, et al. Enrichment methods provide a feasible approach to comprehensive and adequately powered investigations of the brain methylome. Nucleic Acids Res. 2017;epub 25 February 2017.

28. Aberg KA, Dean B, Shabalin AA, Chan RF, Han LKM, Zhao M, et al. Methylome-wide association findings for major depressive disorder overlap in blood and brain and replicate in independent brain samples. Mol Psychiatry. 2018.

29. Aberg KA, Chan RF, van den Oord E. MBD-seq - realities of a misunderstood method for high-quality methylome-wide association studies. Epigenetics. 2020;15(4):431–8.

30. Langmead B, Salzberg SL. Fast gapped-read alignment with Bowtie 2. Nat Methods. 2012;9(4):357–9.

31. Shabalin AA, Hattab MW, Clark SL, Chan RF, Kumar G, Aberg KA, et al. RaMWAS: Fast Methylome-Wide Association Study Pipeline for Enrichment Platforms. Bioinformatics. 2018.

32. van den Oord EJ, Sullivan PF. False discoveries and models for gene discovery. Trends Genet. 2003;19(10):537–42.

33. Shen-Orr SS, Gaujoux R. Computational deconvolution: extracting cell type-specific information from heterogeneous samples. Curr Opin Immunol. 2013;25(5):571–8.

34. Roadmap Epigenomics C, Kundaje A, Meuleman W, Ernst J, Bilenky M, Yen A, et al. Integrative analysis of 111 reference human epigenomes. Nature. 2015;518:317.

35. Kim D, Langmead B, Salzberg SL. HISAT: a fast spliced aligner with low memory requirements. Nat Methods. 2015;12(4):357–60.

36. Pertea M, Pertea GM, Antonescu CM, Chang TC, Mendell JT, Salzberg SL. StringTie enables improved reconstruction of a transcriptome from RNA-seq reads. Nat Biotechnol. 2015;33(3):290–5.

37. Imai K, Keele L, Tingley D. A General Approach to Causal Mediation Analysis. Psychological Methods. 2010;15(4):309–34.

38. Costello EJ, Angold A, Burns B, Stangl D, Tweed D, Erkanli A, et al. The Great Smoky Mountains Study of Youth: Goals, designs, methods, and the prevalence of DSM-III-R disorders. Archives of General Psychiatry. 1996;53:1129–36.

39. Praschberger R, Lowe SA, Malintan NT, Giachello CNG, Patel N, Houlden H, et al. Mutations in Membrin/GOSR2 Reveal Stringent Secretory Pathway Demands of Dendritic Growth and Synaptic Integrity. Cell Rep. 2017;21(1):97–109.

40. Plaud C, Joshi V, Marinello M, Pastre D, Galli T, Curmi PA, et al. Spastin regulates VAMP7-containing vesicles trafficking in cortical neurons. Biochim Biophys Acta Mol Basis Dis. 2017;1863(6):1666–77.

41. Coba MP, Ramaker MJ, Ho EV, Thompson SL, Komiyama NH, Grant SGN, et al. Dlgap1 knockout mice exhibit alterations of the postsynaptic density and selective reductions in sociability. Sci Rep. 2018;8(1):2281.

42. Fan Z, Qian Y, Lu Q, Wang Y, Chang S, Yang L. DLGAP1 and NMDA receptor-associated postsynaptic density protein genes influence executive function in attention deficit hyperactivity disorder. Brain Behav. 2018;8(2):e00914.

43. Stewart SE, Yu D, Scharf JM, Neale BM, Fagerness JA, Mathews CA, et al. Genome-wide association study of obsessive-compulsive disorder. Mol Psychiatry. 2013;18(7):788–98.

44. Guintivano J, Shabalin AA, Chan RF, Rubinow DR, Sullivan PF, Meltzer-Brody S, et al. Test-statistic inflation in methylome-wide association studies. Epigenetics. 2020;15(11):1163–6.

45. Clark SL, Costin BN, Chan RF, Johnson AW, Xie L, Jurmain JL, et al. A Whole Methylome Study of Ethanol Exposure in Brain and Blood: An Exploration of the Utility of Peripheral Blood as Proxy Tissue for Brain in Alcohol Methylation Studies. Alcohol Clin Exp Res. 2018;42(12):2360–8.

46. Hasler R, Preti MG, Meskaldji DE, Prados J, Adouan W, Rodriguez C, et al. Inter-hemispherical asymmetry in default-mode functional connectivity and BAIAP2 gene are associated with anger expression in ADHD adults. Psychiatry Res Neuroimaging. 2017;269:54–61.

47. McKinney B, Ding Y, Lewis DA, Sweet RA. DNA methylation as a putative mechanism for reduced dendritic spine density in the superior temporal gyrus of subjects with schizophrenia. Transl Psychiatry. 2017;7(2):e1032.

48. Kang J, Park H, Kim E. IRSp53/BAIAP2 in dendritic spine development, NMDA receptor regulation, and psychiatric disorders. Neuropharmacology. 2016;100:27–39.

49. Lussier AA, Stepien KA, Neumann SM, Pavlidis P, Kobor MS, Weinberg J. Prenatal alcohol exposure alters steady-state and activated gene expression in the adult rat brain. Alcohol Clin Exp Res. 2015;39(2):251–61.

50. Liu XS, Wu H, Ji X, Stelzer Y, Wu X, Czauderna S, et al. Editing DNA Methylation in the Mammalian Genome. Cell. 2016;167(1):233-47.e17.

