## Supplemental Text for "Dual methylation and hydroxymethylation study of alcohol use disorder"

### Supplemental Material

**Quality control of the brain methylation and hydroxymethylation data**

After read alignment with Bowtie2[1] we performed thorough quality control of samples, reads, and sites[2] using the RaMWAS Bioconducter package[3], which is specifically designed for enrichment-based methylation sequencing studies. In total, both the methylation (mCG) and hydroxymethylation (hmCG) data were obtained from 50 brain autopsies from 50 unique individuals.

Of the generated mCG and hmCG profiles, none were excluded due to failed libraries or sequencing (mainly poor library quality or low number of reads), or because of poor or failed enrichment (peak skewness, unexpected peak size and/or high background levels). The reported sex in the phenotype files were checked for agreement with the overall amount of methylation detected on the sex chromosomes. No sample swaps or errors in the phenotype file were detected. This left 50 samples for further analysis.

For mCG, the mean number of reads for samples used in this study was 54.7 million (SD=3.1 million) of which, on average, 99.3% aligned. Aligned reads were checked for excessive duplicate reads (>3 reads starting at the same location were reset to 1) and reads located in loci where alignment is challenging, determined by an in-silico experiment described elsewhere[2], were excluded. This left an average of 42.7 million (SD=2.8 million) reads per sample (=78.2% of all reads). Akin to filtering SNPs with low minor allele frequency, we excluded rarely methylated sites (average read coverage <0.3). This left 20,823,597 autosomal CpGs for MWAS.

For hmCG, reads and sites were QC'd in a similar manner to the mCG data. The mean number of reads for samples used in this study was 58.1 million (SD=6.2 million) of which, on average, 99.2% aligned. After excluding duplicate reads and reads located in regions that were challenging to align, an average of 54.7 million (SD=6.2 million) reads per sample (=94.1% of all reads) remained. After filtering rare sires, there were 26,153,809 autosomal hydroxymethylation sites for association testing.

**Figure S1 Genomic distribution of methylated and hydroxymethylated sites**
We classified loci as methylated/hydroxymethylated vs. non-methylated/hydroxy, and genomic features as present vs. absent. Using these 2 by 2 tables as input for the colocalization tests described below, we calculated the odds ratios that indicated whether sites in the studied feature were more likely to methylated/hydroxymethylated compared to sites not in this feature. We plotted the odds ratio on the x-axis. Thus, a value of 1, indicated by the dashed lines in the figure, means no enrichment of methylated/hydroxmethylated sites at that feature. The full list of tested features is provided in Supplemental Table S##.

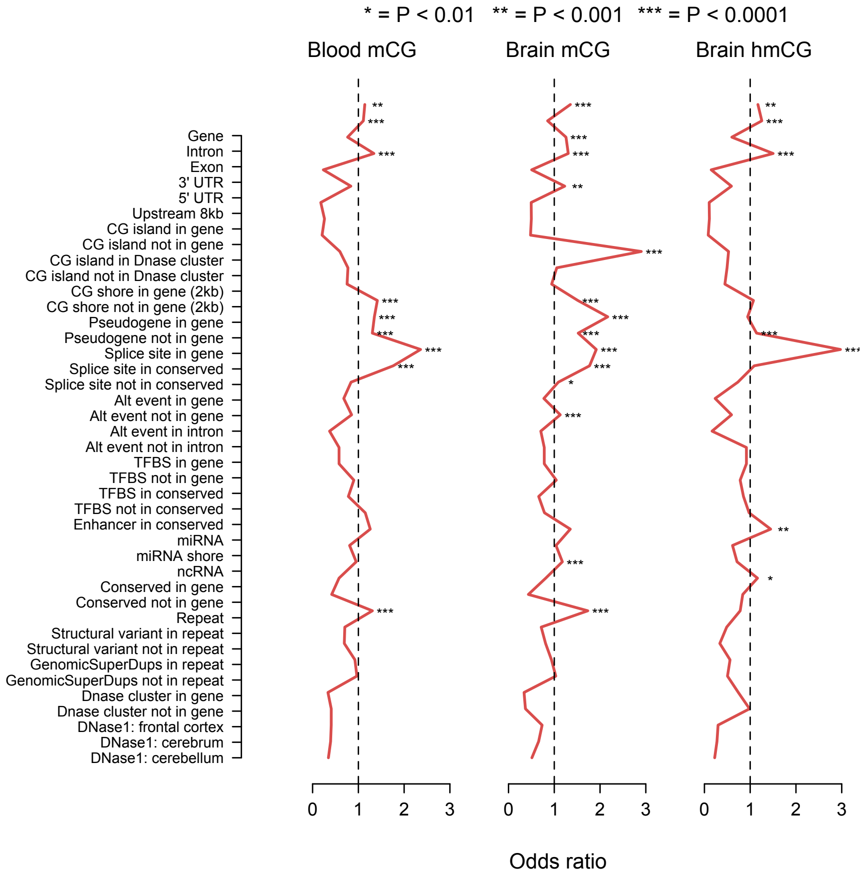

**Cell type specific MWAS**

It is practically not feasible to isolate all common cell types also and perform methylation assays for each type for all study samples. We therefore used a statistical approach to perform cell type specific MWAS. Thus, we first estimated the cell type proportions for each sample using Houseman's method[4,5] and then, following Shen-Orr et al.[6], used the estimated cell type proportions to test the null hypothesis that mean methylation of a given site is equal for AUD cases and controls for each cell type.

In the notation to describe the statistical approach, we indicate constants/parameters with lower case letters and vectors/variables with upper case letters. Furthermore, subscript *i* is used for the *n* subjects and subscript *j* for the *m* methylation sites. Note that whereas analyses in step 1 use all methylation sites for a single subject concurrently, step 2 uses all subjects for a single site concurrently. This is the reason that formulas for step 1 only have subscript *i* (all sites are represented by vectors) and those for step 2 only have subscript *j* (all subjects are represented by vectors).

The amount of methylation in bulk tissue is a weighted sum of the average methylation levels for each cell type with weights being equal to the proportion of cells of each type. For example, if we know the methylation levels for each of the $c = 1..n_c$ cell types we can calculate bulk methylation levels as follows:

$$\bar{Y}_i^{bulk} = \sum_{c=1}^{n_c} p_i^c M_i^c$$

where $m$×1 vector $\bar{Y}_i^{bulk}$ contains the calculated bulk methylation levels for subject *i*, $c$=1..$n_c$ indicates the cell types, $p_i^c$ is the proportion of cells of type *c* for subject *i*, and $M_i^c$ is a $m$×1 vector of cell type *c* specific methylation levels for subject *i*. To avoid performing cell counts for all subjects in the study, the cell type proportions $p_i^c$ are estimated using “reference” methylomes generated with DNA from sorted nuclei/cells using the following regression model:

$$Y_i^{bulk} = \sum_{c=1}^{n_c} p_i^c R^c + E_i$$

where $m$×1 vector $Y_i^{bulk}$ represents the methylation measurements for subject $i$ in bulk tissue, $m$×1 vector $R^c$ contains for each site the mean methylation across the reference methylomes for cell type $c$, and $E_i$ is a $m$×1 vector with residuals. Thus, the methylation levels assayed in bulk tissue for subject $i$ is again a weighted sum of the average methylation levels in the cell type specific methylomes but we replace $M_i^c$ with the reference samples. The weights or cell type proportions are the estimated coefficients of the regression model. The reference methylomes will not perfectly match the true cell type methylation profiles for subject $i$ and reference methylomes may not be available for rare cell types. The model account for that through estimated residuals.

In the second step we use the estimated cell type proportions to fit a regression model that allows us to test the null hypothesis that cell type means are the same in cases and controls ($H_0$:$m_{j(AUD)}^c$ =0):

$$Y_j^{bulk} = \sum_{c=1}^{n_c} m_j^c P^c + \sum_{c=1}^{n_c} m_{j(AUD)}^c (AUD \times P^c) + E_j$$

where $n$×1 vector $Y_j^{bulk}$ are the methylation measurements for site $j$ in bulk, $c$=1..$n_c$ indicate the cell types, $P^c$ is the $n$×1 vector with the proportion of cells of type $c$,$m_j^c$ is the effect of cell type on methylation levels in bulk for site $j$. The $n$×1 vector AUD is coded 1 for cases and 0 for controls where parameter $m_{j(AUD)}^c$ represents the case-control difference for cell type $c$ and site $j$. $E_j$ is a $n$×1 vector with residuals. Thus, the bulk methylation levels for site $j$ are regressed on the cell type proportions $P^c$. This will estimate the mean for each cell type in controls $m_j^c$. The effect of the product AUD× $P^c$, that is zero in controls but $m_{j(AUD)}^c$ in cases, captures the case control difference for cell type *c.*

**Cross-tissue Overlap and Colocalization Testing**

We used circular permutations[7] in the cross-tissue overlap tests to account for dependency between sites. In other words, as the permutations are performed on a per site basis (i.e., individual mCG or hmCG sites), they account for gene size (i.e., genes with more CpGs are more likely to be among the top results in the permutations) and correlations between neighboring sites (e.g., linkage disequilibrium). To perform permutations of overlap between datasets we map the two datasets to each other based on chromosomal location. Next, the P-values for each site are used to cross-classify each mapped marker in the two datasets as being in the top or bottom. Based on the resulting 2 by 2 tables as input, we test the null hypothesis that the enrichment odds ratio equals 1. To perform these tests, we use circular permutations that shifts the mapping of the two datasets by a single random integer in each permutation. This approach to generate the empirical test statistic distribution under the null hypothesis preserves the correlational structure of the data. Multiple thresholds can be specified to define “top findings” (i.e., for our cross-tissue analyses we used the top 0.1%, and 0.5%). To account for this “multiple testing”, the same thresholds are used in the permutations where the test statistic distribution under the null hypothesis is generated from the most significant (combination of) thresholds.

For co-localization analyses such as those involving genomic features and chromatin states, we used a site-based test (i.e., individual mCG or hmCG sites), as these features may not necessarily involve genes. Tests were performed using our R package shiftR (https://cran.rproject.org/web/packages/shiftR/index.html) that performs circular permutations through bitwise operations. shiftR was used as these bitwise operations are faster for CpG based tests. We first mapped the CpGs of MWAS to the other data set based on chromosomal location. Next, the state values are used to cross-classify each mapped site in the two data sets as being in the top or bottom. Based on the resulting 2 by 2 tables as input, shiftR tests uses Cramér's V to test the null hypothesis that the enrichment odds ratio equals one. To perform these tests, shiftR uses circular permutations. Specifically, through these fast, bitwise operations, it shifts the mapping of the two data sets by a single random integer in each permutation. If multiple thresholds are specified to define "top findings", the same thresholds are used in the permutations where the empirical test statistic distribution under the null hypothesis is generated from most significant (combination of) thresholds.

For the co-localization analyses, we tested whether the overlap across blood and brain colocalized with basic genomic feature tracks that were downloaded from the UCSC Browser Annotation Database and Roadmap Epigenomics Project chromHMM 15-state chromatin state consensus tracks[8]. All co-localization analyses were performed using shiftR using 100,000 permutations. For the chromatin state colocalization analyses, we used Quiescent/Low as the reference state. To study overlap between the 15 histone states, we used the E073 track from the Dorsolateral Prefrontal Cortex for the enrichment testing.

**Quality control of the transcript expression data**

After read alignment with HiSat2[9] there was an average of 63.4 million reads per sample (S.D. = 6.3 million). Reads were assembled into transcripts and quantified using StringTie[10]. Transcripts were then compared against known transcripts and genes using gffcompare utility of StringTie[10]. Further quality control was conducted using custom R scripts. In total, RNA-seq data was obtained from the same 50 samples for which we generated methylation and hydroxymethylation.

After assembly, there were 227,442 unique transcripts of which 195,811 (86.1%) were located in known transcripts. After removing unknown transcripts and retaining those on the autosomal chromosomes, 188,192 transcripts remained. Following similar quality control procedures to what was used in Gtex (cite), we retained transcripts that had TPM > 0.1 in at least 20% of the samples, greater than or equal to 5 reads per sample, and average TPM > 1. This left 40,090 high quality transcripts for analysis. Transcripts were then logged plus one and quantile normalized.

**Replication Sample: Great Smokey Mountain Study**

*Participants*

The Great Smoky Mountain Study (GSMS) is an ongoing, prospective longitudinal, representative study of children in 11 predominantly-rural counties in the southeast United States, which began in 1993[11]. Three cohorts of children, age 9 to 13 years, were recruited resulting in N=1,420 participants. Details about the GSMS design, recruitment and data collection are published elsewhere[11]. As part of a larger methylation study on childhood and young adult exposures, 525 participants were selected that were 9-21 years of age and had a bloodspot taken at the time of assessment available. Of the 525 participants, 412 had bloodspots included from additional older time points (age 21 years or older). Only data from participants that were 18 years of age or older were used in this study to be comparable to the discovery sample. Participants provided their informed consent and the current study was approved by Institutional Review Boards at Duke University and Virginia Commonwealth University.

Participant substance involvement was assessed during an in-person interview. At each interview assessment the participant completed a full structured clinical interview about substance involvement using the Young Adult Psychiatric Assessment (YAPA)[12]. The substance use module of the YAPA included assessments of DSM-IV abuse and dependence. Although

DSM-5 alcohol use disorder (AUD) symptoms of craving and withdrawal were not part of the DSM-IV abuse or dependence diagnostic criteria, these data have been collected since the start of GSMS is 1993. Further details about the study sample, and the demographic and clinical characteristics of participants used for the present study are in Table S17.

*Assaying the Methylome*

The same MBD-seq approach, which was previously used to generate mCG data for the discovery blood samples [13] was also used to generate mCG data for the independent replication samples. Specifically, we used an optimized protocol for the MBD-seq approach [14,15] that achieves near-complete coverage of the 28 million possible mCG sites at a cost comparable to commonly used methylation arrays that assay only 2-3% of all mCG sites [16]. Briefly, genomic DNA was sheared into 150 bp fragments using the Covaris™ E220 focused ultrasonicator system. We performed enrichment with MethylMiner™ (Invitrogen) to capture the methylated fraction of the genome. Next, dual-indexed sequencing library for each methylation capture was prepared using the Accel-NGS® 2S Plus DNA Library Kit (Swift Biosciences) and sequenced on a NextSeq500 instrument (Illumina). The sequence reads were aligned to the human reference genome (hg19/GRCh37) using Bowtie2[1]. Following quality control, libraries were pooled in equal molarities and sequenced with 75 cycles (i.e., 1 x 75 base pair reads) on a NextSeq500 instrument (Illumina).

*Data Processing and Methylation Score Calculation*

As in the discovery, data quality control and analyses were performed in RaMWAS[17] and followed the same steps used for the discovery sample. Methylation scores were calculated by estimating the number of fragments covering the CpG using a non-parametric estimate of the fragment size distribution. These scores provide a quantitative methylation measure for each individual at that specific site.

*Methylome-wide Association Study in Whole Blood*

The methylome-wide association study (MWAS) in whole blood was performed using multiple regression analyses with four sets of covariates. First, we regressed out assay-related variables (i.e., potential technical artifacts) such as sample batches and peak[17]. Second, we regressed out biological sex, age, age squared, race and regular cigarette use. Third, to avoid false positives due to cell type heterogeneity in whole blood, we regressed out cell type proportions as estimated from the methylation data[4]. Fourth, principle component analysis (PCA) was used to capture any remaining unmeasured sources of variation. One principal component was selected based on the Scree test. A false discovery rate (FDR) of 10% (q-value < 0.1) was used to classify sites as differentially methylated.

*Cell-type Specific MWAS*

We used the same epigenomic deconvolution approach used in the discovery (described above) to perform cell-type specific MWAS for the major nucleated cell types found in blood: granulocytes (CD15+), T-cells (CD3+), B-cells (CD19+) and monocytes (CD14+). Assay-related variables, demographic variables (sex, age, age squared, race, regular cigarette use), and one principal component were included as covariates in the cell-type specific MWAS. Cell-type proportions were also included as main effects in cell-type specific MWAS as the epigenomic deconvolution is essentially an interaction model[18].

**Block-based Association Testing**

To potentially reduce the multiple testing burden in the brain association analyses, the sites were adaptively combined by collapsing highly inter-correlated coverage estimates at adjacent sites into a single mean coverage estimate [19]. This was done separately for mCG and hmCG. This resulted in 10,082,773 "blocks" for mCG and 17,755,251 for hmCG, which were then tested for association with AUD. The brain mCG and hmCG block association testing was conducted separately for mCG and hmCG, and included measured technical variables, sex, age at death, post-mortem interval, estimated cell type proportions, and one principal component.

**Figure S2 Quantile-Quantile plot for block-based association testing in brain mCG**
The observed p-values (red dots), on a -log10 scale, are plotted against their expected values (grey main diagonal line) under the null hypothesis assuming none of the sites have an effect. Orange lines indicate the 95% confidence bands (CI). A deviation of the observed p-values from the main diagonal indicates that there are sites associated with AUD. Coefficient lambda ($\lambda$) will be close to one if the vast majority of sites behave as expected under the null hypothesis.

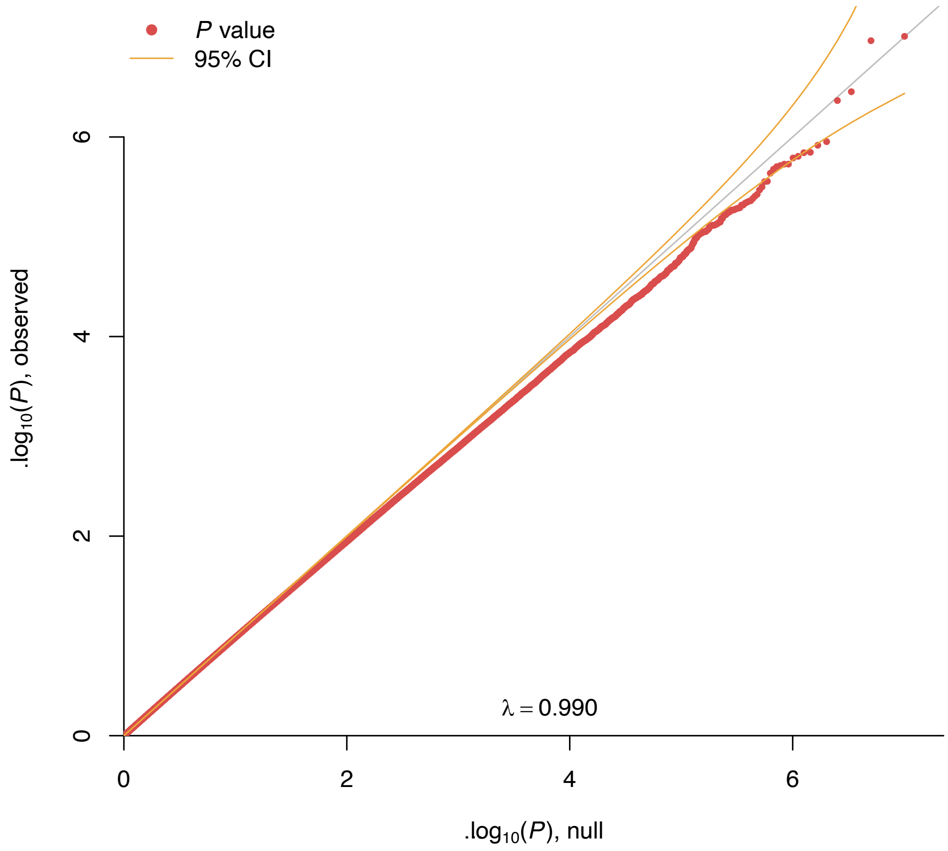

**Figure S3 Quantile-Quantile plot for block-based association testing in brain hmCG**
The observed p-values (red dots), on a -log10 scale, are plotted against their expected values (grey main diagonal line) under the null hypothesis assuming none of the sites have an effect. Orange lines indicate the 95% confidence bands (CI). A deviation of the observed p-values from the main diagonal indicates that there are sites associated with AUD. Coefficient lambda ($\lambda$) will be close to one if the vast majority of sites behave as expected under the null hypothesis.

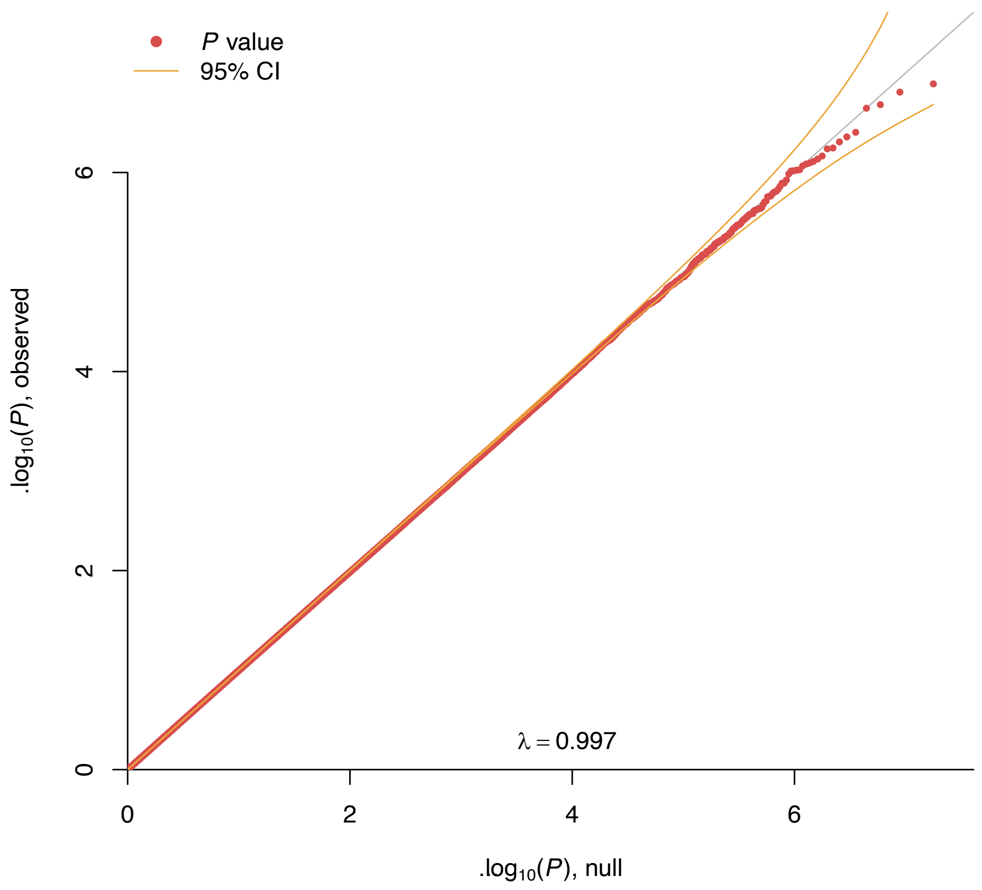

### References

1. Langmead B, Salzberg SL. Fast gapped-read alignment with Bowtie 2. *Nat Methods.* 2012;9(4):357-359.
2. Aberg KA, McClay JL, Nerella S, et al. MBD-seq as a cost-effective approach for methylome-wide association studies: demonstration in 1500 case--control samples. *Epigenomics.* 2012;4(6):605-621.
3. Shabalin AA, Hattab MW, Clark SL, et al. RaMWAS: Fast Methylome-Wide Association Study Pipeline for Enrichment Platforms. *Bioinformatics.* 2018.
4. Houseman EA, Accomando WP, Koestler DC, et al. DNA methylation arrays as surrogate measures of cell mixture distribution. *BMC Bioinformatics.* 2012;13:86.
5. Koestler DC, Christensen B, Karagas MR, et al. Blood-based profiles of DNA methylation predict the underlying distribution of cell types: a validation analysis. *Epigenetics.* 2013;8(8):816-826.
6. Shen-Orr SS, Tibshirani R, Khatri P, et al. Cell type-specific gene expression differences in complex tissues. *Nat Methods.* 2010;7(4):287-289.
7. Cabrera CP, Navarro P, Huffman JE, et al. Uncovering networks from genome-wide association studies via circular genomic permutation. *G3 (Bethesda).* 2012;2(9):1067-1075.
8. Roadmap Epigenomics C, Kundaje A, Meuleman W, et al. Integrative analysis of 111 reference human epigenomes. *Nature.* 2015;518:317.
9. Kim D, Langmead B, Salzberg SL. HISAT: a fast spliced aligner with low memory requirements. *Nat Methods.* 2015;12(4):357-360.
10. Pertea M, Pertea GM, Antonescu CM, Chang TC, Mendell JT, Salzberg SL. StringTie enables improved reconstruction of a transcriptome from RNA-seq reads. *Nat Biotechnol.* 2015;33(3):290-295.
11. Costello EJ, Angold A, Burns B, et al. The Great Smoky Mountains Study of Youth: Goals, designs, methods, and the prevalence of DSM-III-R disorders. *Archives of General Psychiatry.* 1996;53:1129-1136.
12. Angold A, Cox A, Prendergast M, et al. *The Young Adult Psychiatric Assessment (YAPA).* Durham, NC: Duke University Medical Center;1999.
13. Aberg KA, Dean B, Shabalin AA, et al. Methylome-wide association findings for major depressive disorder overlap in blood and brain and replicate in independent brain samples. *Mol Psychiatry.* 2018.
14. Aberg KA, Chan RF, Shabalin AA, et al. A MBD-seq protocol for large-scale methylome-wide studies with (very) low amounts of DNA. *Epigenetics.* 2017:0.
15. Chan RF, Shabalin AA, Xie LY, et al. Enrichment methods provide a feasible approach to comprehensive and adequately powered investigations of the brain methylome. *Nucleic Acids Res.* 2017;epub 25 February 2017.
16. Aberg KA, Chan RF, van den Oord E. MBD-seq - realities of a misunderstood method for high-quality methylome-wide association studies. *Epigenetics.* 2020;15(4):431-438.
17. Shabalin AA, Hattab MW, Clark SL, et al. RaMWAS: fast methylome-wide association study pipeline for enrichment platforms. *Bioinformatics.* 2018.
18. Shen-Orr SS, Gaujoux R. Computational deconvolution: extracting cell type-specific information from heterogeneous samples. *Curr Opin Immunol.* 2013;25(5):571-578.
19. Aberg K, Khachane AN, Rudolf G, et al. Methylome-wide comparison of human genomic DNA extracted from whole blood and from EBV-transformed lymphocyte cell lines. *Eur J Hum Genet.* 2012;20(9):953-955.
